# Tobacco and Nicotine Use in the United States Army Trainee Soldier’s Environment

**DOI:** 10.1101/2021.07.16.21260650

**Authors:** Adam Edward Lang, Aleksandra Yakhkind, Elizabeth Prom-Wormley

**Author notes:** Correspondence Author: Adam Edward Lang |, McDonald Army Health Center, 576 Jefferson Avenue, Fort Eustis, Virginia 23604. The views expressed in this publication are those of the author and do not necessarily reflect the official policy of the Department of Defense, Department of the Army, U.S. Army Medical Department, or the U.S. Government.

## Abstract

Tobacco and nicotine use rates within the United States military population are high, and rates among young trainee soldiers are not improving over time. Trainee soldiers are heavily influenced by role models in their environment, and nicotine product use rates in this environment has not previously been evaluated. This study found that over 30% of non-trainee U.S. Army soldiers, known as cadre, within the training environment use nicotine products. For the young trainee soldier, this creates an initial Army experience that continues to foster a culture of tobacco and nicotine use. We urge changes in Army tobacco and nicotine policy to protect the vulnerable trainee soldier population.

## Introduction

Rates of tobacco and nicotine product use in United States (US) Army trainees in 2019 were shown to be over 35%, which was significantly higher compared to those aged 18-24 in the general US population that same year (1, 2). One promising strategy to reduce the rates of tobacco and nicotine use in soldiers is the implementation of policies restricting their use. Specifically, the use of these products is not allowed in the first 10-week phase of Army training (basic combat training, BCT). Rates of nicotine product use in trainees who smoked prior to BCT decreased by 31.5% after this stage of training (1). Nicotine use is allowed in the second phase (advanced individual training, AIT). This shift in policy towards nicotine use has allowed trainees who used prior to BCT to reinitiate early in a cessation attempt. It also puts trainees who never used nicotine products in an environment where peer nicotine product use is allowed and accepted.

In 2020, a nicotine-free policy was implemented throughout an AIT brigade on Fort Eustis in Virginia, where training spans on average three to six months, in order to encourage nicotine abstinence and extend the trainee nicotine-free period. AIT trainees were subject to this policy, but cadre were not. Cadre include all non-trainee active duty service members such as drill sergeants, course instructors, and those in command positions. Cadre hold enormous influence over trainees through constant daily interactions. Consequently, AIT trainees may be exposed to inconsistent messaging regarding social norms related to tobacco and nicotine use in the Army. Our study sought to identify prevalence of tobacco and nicotine use among cadre to understand the potential degree of trainee exposure to tobacco and nicotine during AIT.

## Methods

Data were collected using a confidential online survey between January 08-25, 2020, just prior to the implementation of the AIT nicotine-free policy discussed above. All cadre in an AIT training brigade on Fort Eustis (N = 757) received an email with an invitation to participate in the survey as well as contact information for the tobacco treatment clinic to assist those interested in receiving treatment for tobacco and nicotine dependence. Four hundred and thirteen (N = 413) individuals participated in the survey (54.6% response rate). The study was deemed exempt from review by the Clinical Investigation Department at Naval Medical Center Portsmouth. Statistical analyses were conducted using R, version 3.6.1 (3).

## Results

One hundred and ninety (46.0%) cadre participants engaged in any lifetime nicotine use, 159 (38.4%) used prior to joining the military, and 126 (30.8%) were current nicotine users. Most current users (74.0%) have used nicotine for seven years or more and 38.6% had been using for 15 years or more. Further, 51.1% of current users engaged in nicotine use six or more times per day. The most common product used was smokeless tobacco (40.6%), followed by conventional cigarette use (36.6%). Of current users, 20.5% engaged in dual or polyproduct use. The most common combination of use was combustible cigarette and smokeless tobacco (45.8%), followed by combustible cigarette and electronic cigarette use (20.8%, Table 1).

**Table 1.** Summary Statistics.

| **Variable** | **N** | **%** |
| --- | --- | --- |
| **Gender** |  |  |
| Man | 370 | 89.6 |
| Woman | 34 | 8.2 |
| Other | 9 | 2.2 |
| Total | 413 | 100.0 |
| **Age** |  |  |
| 18-29 | 65 | 15.7 |
| 30-39 | 247 | 59.8 |
| 40 and over | 101 | 24.5 |
| Total | 413 | 100.0 |
| **Any Lifetime Nicotine Use** |  |  |
| No | 223 | 54.0 |
| Yes | 190 | 46.0 |
| Total | 413 | 100.0 |
| **Nicotine Use Prior to the Military** |  |  |
| No | 254 | 61.6 |
| Yes | 159 | 38.4 |
| Total | 413 | 100.0 |
| **Current Nicotine Use** |  |  |
| No | 286 | 69.2 |
| Yes | 127 | 30.8 |
| Total | 417 | 100.0 |
| **Length of Use (in Years)** |  |  |
| Less than 1 | 9 | 7.1 |
| 1-3 | 12 | 9.4 |
| 4-6 | 12 | 9.4 |
| 7-10 | 22 | 17.3 |
| 11-15 | 23 | 18.1 |
| 15 and over | 49 | 38.6 |
| Total | 127 | 99.9 |
| **Frequency of Daily Use** |  |  |
| Less than 1 time/day | 8 | 6.3 |
| 1-5 times/day | 54 | 42.5 |
| 6-10 times/day | 36 | 28.3 |
| More than 10 times/day | 29 | 22.8 |
| Total | 127 | 99.9 |
| **Poly-product Use** |  |  |
| 1 product | 101 | 79.5 |
| 2 products | 24 | 18.9 |
| 3 or more products | 2 | 1.6 |
| Total | 127 | 100.0 |
| **Single Product-Specific Use** |  |  |
| Electronic Cigarette | 20 | 19.8 |
| Conventional Cigarette | 37 | 36.6 |
| Cigar | 3 | 3.0 |
| Smokeless | 41 | 40.6 |
| Dissolvable | 0 | 0.0 |
| Pipe | 0 | 0.0 |
| Total | 101 | 100.0 |
| **Dual Use** |  |  |
| Cigarette/ Smokeless | 11 | 45.8 |
| Cigarette/ Cigar | 5 | 20.8 |
| Cigarette/ Electronic Cigarette | 5 | 20.8 |
| Cigarette/Pipe | 1 | 4.2 |
| Cigar/ Pipe | 1 | 4.2 |
| Electronic Cigarette/ Dissolvable | 1 | 4.2 |
| Total | 24 | 100.0 |
| **Current Desire to Quit** |  |  |
| No | 89 | 70.1 |
| Yes | 38 | 29.9 |
| Total | 127 | 100.0 |
| **Desire to Quit within a Year** |  |  |
| No | 69 | 54.3 |
| Yes | 58 | 45.7 |
| Total | 127 | 100.0 |

At the time of the survey, 29.9% of current users were interested in quitting. The proportion of interest to quit increased to 45.7% when participants were asked whether they wanted to quit within a year.

## Discussion

Cadre tobacco and nicotine use in the military training setting has not previously been evaluated. The results in this study indicate that almost one-third of cadre are current users and that over half engage in a high frequency of nicotine use (6 or more times per day). Consequently, tobacco and nicotine use in cadre is widespread and is expected to be a factor in trainee tobacco and nicotine use. In prior studies, young Air Force trainees as well as civilian adolescents report their tobacco use to be heavily influenced by cadre or role model tobacco use, peer tobacco use, and the individual’s perception of social norms (4, 5). Of concern in our study is the relatively small change in prevalence of cadre nicotine use from before they joined the military until the present time, along with the high percentage of cadre using for greater than 15 years. Given the interest in quitting among cadre, a focus on policy change and appropriate treatment of tobacco and nicotine dependence in soldiers of every age and rank must be a priority for the United States Army. Interventions targeting seasoned soldiers would prevent the downstream effect of influencing use in the trainee soldier. This influence, which likely results in de novo, reinitiated, continued, and multiproduct use, is a major part of the cycle contributing to systematic use within the Army. Interventions are essential to prevent poor health outcomes in military, Veterans Affairs, and civilian populations, and the degradation of US Army readiness.

Study limitations include the potential for recall bias of nicotine use prior to joining the military, absence of objective measurements of nicotine use, self-selection bias, inability to compare demographics of those who responded to survey non-responders, an underrepresentation of women compared to the military average, lack of differentiation between non-nicotine and nicotine e-cigarettes on the survey, and a population of soldiers only within the aviation field.

Future AIT nicotine-free policies must include cadre in order to be maximally effective. The Army trainee population is highly vulnerable. Policies to protect them are realistic in the highly controlled military setting, and lack of action allows the harmful cycle of military tobacco and nicotine use to continue.

## Data Availability

The data that support the findings of this study are available from the corresponding author, AEL, upon reasonable request.

## Abbreviations

AIT: (Advanced Individual Training)
BCT: (Basic Combat Training
US: (United States)

